# Cohort profile: The Cancer Risk Associated with the Body Art of Tattooing (CRABAT) study

**DOI:** 10.1101/2024.10.28.24316120

**Authors:** Bayan Hosseini, Rachel McCarty, Marie Zins, Marcel Goldberg, Céline Ribet, Ines Schreiver, Khaled Ezzedine, Joachim Schüz, Milena Foerster

## Abstract

Tattooing, involving the injection of pigments into the skin, has become increasingly popular, with up to 40% of individuals under 40 years old tattooed in high-income countries. Despite regulatory measures, tattoo inks may contain hazardous substances such as polycyclic aromatic hydrocarbons, primary aromatic amines, and metallic impurities, many of which are classified as carcinogens. Research on systemic health risks, particularly cancer, associated with intradermal ink exposure remains limited. Complications like contact allergies and inflammatory skin reactions are more frequently reported among tattooed individuals. However, cancer risks from tattooing, especially for internal cancers like lymphoma, are challenging to assess. Existing case-control studies indicate mixed findings regarding hematologic cancers, with one large study reporting a 20% increased lymphoma risk among tattooed individuals in the first two years post-tattooing.

Here, we present the Cancer Risk Associated with the Body Art of Tattooing (CRABAT) study, that is, to our knowledge, the first larger study that prospectively investigates tattoo-related cancer risks. CRABAT follows up over 110,000 participants for long-term health effects within the French Constances cohort with objective cancer data linkage. Of ∼13,000 participants that were tattooed in 2020, detailed tattoo exposure assessment via a validated tattoo exposure questionnaire was conducted in 2023 (response rate >60%). With its robust dataset through linkage to the Constances variable pool, CRABAT enables the analysis of exposure-response relationships, and addresses potential confounders such as sociodemographic and lifestyle factors, and alternative pathways such as tattoo-related infections. Body word count: 2498 words

## Introduction

Tattooing involves the injection of organic or inorganic pigment particles diluted in a carrier liquid into the dermis. Since the 1990s tattoos have gained extreme popularity with up to 40% in under 40-year-olds in high income countries being tattooed (1). Still few people are aware that despite existing regulations in Europe, tattoo inks may contain hazardous chemicals, such as polycyclic aromatic hydrocarbons (PAHs) bound to carbon black particles, soluble primary aromatic amines (PAAs) in brightly coloured inks, and metallic impurities such as nickel, and chromium, in all kinds of ink (2-4). Many of these substances have been classified by the International Agency for Research on Cancer (IARC) as class 1, 2A or 2B carcinogens, based on respiratory or oral exposure and/or topical application (5), (6, 7). The potential health risks of the complex intradermal tattoo ink exposure leading to lymphatic accumulation of pigment particles remain under-researched.(8, 9) However, with more tattooed individuals, tattoo related complications such as contact allergies or inflammatory skin conditions (e.g. granuloma or pseudo lymphoma) become more frequent (9, 10). Only one published case-control study has investigated skin cancer risk associated with tattooing, though the study was limited by a small number of tattooed individuals (11). Systemic long-term health risks like lymphoma or solid cancers of inner organs, that would not appear on the tattoo itself, are more difficult to detect. Three published case-control studies have investigated potential hematologic cancer risk associated with tattooing (12-14). The findings of these studies were mixed with two observing no increased risk of overall hematologic cancers associated with tattooing, though there was some evidence of increased risk of certain B cell lymphoma subtypes. (13, 14) Two of these studies were limited by small sample sizes within cancer subtype strata and low tattoo prevalence. The third study with over 1,300 cases and 4,000 controls using Swedish population registry data, found a 20% increased risk for overall lymphoma in tattooed compared with non-tattooed individuals, with the highest risk in the 0-2 years following a first tattoo (12). Limitations of all three studies included the potential for response bias from the case-control design, limited tattooing exposure data and not controlling for tattoo related infections as alternative pathway in between tattooing and lymphoma.

To overcome these shortcomings, we launched the Cancer Risk Associated with the Body Art of Tattooing (CRABAT) study, which is embedded in the prospective lifetime cohort Constances (Cohorte des consultants des Centres d’examens de santé) and provides a long-term follow-up of cancer (and other chronic conditions) incidence, allowing causal conclusions to be drawn.

## Methods

The Constances study is a large ongoing population-based cohort study across France whose primary aim is to provide an exhaustive research infrastructure for diverse epidemiological research projects within the cohort population and whose protocol is published (15). In brief, over 220,000 volunteers, age 18-to 69-years-old underwent baseline investigations from late 2012 to 2020 (16). The baseline data was collected through self-administered questionnaires that collected information on demographics and detailed medical histories across 20 different departments of France. Constances’ robust design provides a valuable resource for investigating a wide range of health outcomes and their determinants including cancers.

The CRABAT study is a nested cohort study within Constances. This study applied a two-stage process to assess tattoo exposure. First, basic exposure information was collected via the screening questions, “Do you have a tattoo?” and a respective tattoo surface (more or less than 1 hand surface) during the annual follow-up in 2020/21. All respondents to these questions were subsequently enrolled, with tattooed participants constituting the exposed group and non-tattooed participants serving as controls. In the second phase, conducted between July and December 2023, a self-administered tattoo exposure questionnaire (EpiTAT) was distributed to all individuals with tattoos, either online or on paper (17). In April 2024 the CRABAT database was created, and the tattoo exposure data were merged with selected variables from the Constances data for all participants with known tattoo exposure status.

Follow-ups of the Constances cohort are performed annually, via self-administered questionnaires, either using paper-based or web-based questionnaires. The participants have been invited for heath examination every 5 years. Linkage to national social health administrative databases is continuous since 01 January 2007, updated with a 2-year reporting delay. A follow-up tattoo exposure assessment is planned for 2030.

Exposure data were collected using the validated tattoo exposure questionnaire, EpiTAT, which was specifically designed to capture various visual and contextual factors associated with tattoo exposure and that is freely accessible (17). Visual factors included the measurement of tattooed body surface in terms of “number of hand surfaces,” both overall and by anatomical location, as well as tattoo colours, degree of tattoo filling (illustrated by visual examples representing 5%, 25%, 50%, 75%, and 100% of the tattoo fill), and tattoo style. Contextual factors included the age of tattoo acquisition by 5-year age groups, the circumstances of tattoo acquisition, and whether tattoos were obtained outside of France, with respective countries recorded if applicable. Additionally, the questionnaire assessed tattoo complications, whether associated with issues such as poor wound healing or aftercare, and information related to tattoo removal.

Using the collected exposure data, different exposure metrics were derived, including total tattooed body surface area, overall and stratified for anatomical location in hand palms and cm^2^ (hand surface area estimated in cm^2^ according to Sacco et.al) (18). Subsequently, total ink exposure was estimated by adjusting tattooed surface area by multiplication with the proportion of tattoo filling (e.g. in case of 25% filling, the tattooed body surface was multiplied with 0.25).

### Additional self-reported data

Socio-demographic and lifestyle data include age in years, sex at birth (male/ female), education and income level, marital status, origin, household average net income, and occupational status, body mass index (BMI), alcohol consumption, cigarette smoking, e-cigarette consumption, and cannabis consumption (Table 1).. Supplementary table 1 shows additional collected sociodemographic, lifestyle, environmental and medical variables. Furthermore, data on co-exposures including metals, fumes, solvents, pesticides and chemical cleaning products can be included in cancer risk analyses. In a recent analyses of tattooing associated hepatitis risk, this excellent data allowed us to adjust analyses for sex amongst men, multiple sexual partners, condom use, alcohol use, educational status (*manuscript under review*). Furthermore, changes in lifestyle, and other confounders over time can be considered through their repeated assessment during regular cohort follow-ups.

**Table 1:** Sociodemographic characteristics of the study population by tattoo status, by sex.

|  | Overall<br>(n=113,222) |  | <sup>a</sup> P-value | Tattooed<br>(n=13,135) |  | Non-tattooed<br>(n=100,087) |  |
| --- | --- | --- | --- | --- | --- | --- | --- |
|  | Tattooed<br>(n=13,135) | Non-tattooed<br>(n= 100,087) |  | Male<br>(n= 4,889; 37.2%) | Female<br>(n= 8,246; 62.8%) | Male<br>(n= 46,176; 46.1%) | Female<br>(n= 53,911; 53.9%) |
| **Age (year)** | 57.1 (13.4) | 55.9 (13.5) |  | 51.7 (11.7) | 46.3 (11.1) | 57.6 (13.4) | 57.4 (13.2) |
| **Mean (SD)** |  |  |  |  |  |  |  |
|  | n (col% /row%) | n (col% /row%) |  | n (col% /row%) | n (col% /row%) | n (col% /row%) | n (col% /row%) |
| **Age (10 year-interval)** |  |  | <0.001 |  |  |  |  |
| <45 yrs. | 5,434 (41.4, 21.3) | 20,105 (20.1, 78.7) |  | 1,500 (30.7/ 27.6) | 3,934 (47.7/ 72.4) | 9,371 (20.3/ 46.6) | 10,734 (19.9/ 53.4) |
| 45-54 yrs. | 3,807 (29.0, 15.0) | 21,629 (21.6, 85.0) |  | 1,401 (28.7/ 36.8) | 2,406 (29.2/ 63.2) | 9,891 (21.4/ 45.7) | 11,738 (21.8/ 54.3) |
| 55-64 yrs. | 2,531 (19.3, 10.5) | 21,521 (21.5, 89.5) |  | 1,207 (24.7/47.7) | 1,324 (16.1/ 52.3) | 9,489 (20.5/ 44.1) | 12,032 (22.3/ 55.9) |
| 65-74 yrs. | 1,198 (9.1, 4.2) | 27,018 (27.0, 95.8) |  | 687 (14.1/ 55.3) | 511 (6.2/ 42.7) | 12,566 (27.2/ 46.5) | 14,452 (26.8/ 53.5) |
| >=75 yrs. | 165 (1.2, 1.7) | 9,814 (9.8, 98.3) |  | 94 (1.8/ 57.0) | 71 (0.8/ 43.0) | 4,859 (10.6/ 49.5) | 4,955 (9.2/ 50.5) |
| **Tobacco smoking** |  |  | <0.001 |  |  |  |  |
| Never | 4,425 (33.7, 8.1) | 50,204 (50.2, 91.9) |  | 1,365 (27.9/ 30.8) | 3,060 (37.1/ 69.2) | 20,809 (45.1/ 41.4) | 29,395 (54.5/ 58.6) |
| Former | 4,717 (35.9, 12.2) | 33,849 (33.8, 87.8) |  | 2,003 (41.0/ 42.5) | 2,714 (32.9/ 57.5) | 17,635 (38.2/ 52.1) | 16,214 (30.1/ 47.9) |
| Current | 3,459 (26.3, 21.9) | 12,328 (12.3, 78.1) |  | 1,302 (26.6/ 37.6) | 2,157 (26.2/ 62.4) | 6,001 (13.0/ 48.7) | 6,327 (11.7/ 51.3) |
| Missing | 534 (4.1, 12.6) | 3,706 (3.7, 87.4) |  | 219 (4.5/ 41.0) | 315 (3.8/ 59.0) | 1,731 (3.7/ 46.7) | 1,975 (3.7/ 53.3) |
| **Alcohol consumption (Audit score)** |  |  | <0.001 |  |  |  |  |
| None | 267 (2.0, 7.8) | 3,160 (3.2, 92.2) |  | 72 (1.5/ 27.0) | 195 (2.4/ 73.0) | 941 (2.0/ 29.8) | 2,219 (4.1/ 70.2) |
| Abuse | 2,647 (20.3, 15.1) | 14,898 (14.9, 84.9) |  | 1,102 (22.5/ 41.6) | 1,545 (18.7/ 58.4) | 8,538 (18.6/ 57.3) | 6,360 (11.8/ 42.7) |
| Dependence | 806 (6.1, 18.4) | 3,573 (3.6, 81.6) |  | 466 (9.5/ 57.8) | 340 (4.1/ 42.2) | 2,514 (5.4/ 70.4) | 1,059 (2.0/ 29.6) |
| No risk consumption | 8,529 (64.9, 10.5) | 72,405 (72.3, 89.5) |  | 2,990 (61.2/ 35.1) | 5,539 (67.2/ 64.9) | 31,992 (69.3/ 44.2) | 40,413 (75.0/ 55.8) |
| Unknown/ missing | 886 (6.7, 12.8) | 6,051 (6.0, 87.2) |  | 259 (5.3/ 29.2) | 627 (7.6/ 70.8) | 2,191 (4.7/ 36.2) | 3,860 (7.1/ 63.8) |
| **E-cigarette** |  |  | <0.001 |  |  |  |  |
| Never | 8,038 (61.2, 11.2) | 63,506 (63.5, 88.8) |  | 2,948 (60.3/ 36.7) | 5,090 (61.7/ 63.3) | 29,637 (64.1/ 46.7) | 33,869 (62.8/ 53.3) |
| Ever | 1,959 (14.9, 27.0) | 5,305 (5.3, 73.0) |  | 781 (16.0/ 39.9) | 1,178 (14.3/ 60.1) | 2,754 (6.0/ 51.9) | 2,551 (4.7/ 48.1) |
| Missing | 3,138 (23.9, 9.1) | 31,276 (31.2, 90.9) |  | 1,160 (23.7/ 37.0) | 1,978 (24.0/ 63.0) | 13,785 (29.9/ 44.1) | 17,491 (32.5/ 55.9) |
| **Cannabis consumption** |  |  | <0.001 |  |  |  |  |
| Never | 5,395 (42.1/ 7.7) | 64,575 (64.5, 92.3) |  | 1,739 (35.6/ 32.2) | 3,656 (44.3/ 67.8) | 27,601 (59.8/ 42.7) | 36,974 (68.6/ 57.3) |
| Ever | 7,278 (55.4/ 19.3) | 30,511 (30.5, 80.7) |  | 2,951 (60.3/ 40.5) | 4,327 (52.5/ 59.4) | 16,696 (36.1/ 54.7) | 13,815 (25.6/ 45.3) |
| Unknown/ missing | 462 (3.5/ 8.5) | 5,001 (5.0, 91.5) |  | 199 (4.1/ 43.1) | 263 (3.2/ 56.9) | 1,879 (4.1/ 37.6) | 3,122 (5.8/ 62.4) |
| **Body Mass Index kg/M<sup>2</sup>** |  |  | <0.001 |  |  |  |  |
| <18.5 (underweight) | 423 (3.2, 13.9) | 2,618 (2.6, 86.1) |  | 49 (1.0/ 11.6) | 374 (4.5/ 88.4) | 482 (1.0/ 18.4) | 2,136 (4.0/ 81.6) |
| 18.5-24.9 (healthy range) | 7,404 (56.4, 11.6) | 56,583 (56.5, 88.4) |  | 2,317 (47.5/ 31.3) | 5,087 (61.7/ 68.7) | 22,968 (49.7/ 40.6) | 33,615 (62.4/ 59.4) |
| 25-29.9 (overweight) | 3,704 (28.2, 10.9) | 30,128 (30.1, 89.1) |  | 1,884 (38.5/ 50.9) | 1,820 (22.2/ 49.1) | 17,584 (38.1/ 58.4) | 12,544 (23.2/ 41.6) |
| 30-39.9 (obesity) | 1,501 (11.4, 12.9) | 10,165 (10.2, 87.1) |  | 613 (12.5/ 40.8) | 888 (10.7/ 59.2) | 4,965 (10.8/ 48.8) | 5,200 (9.6/ 51.2) |
| >40 (severe obesity) | 103 (0.8, 14.8) | 593 (0.6, 85.2) |  | 26 (0.5/ 25.2) | 77 (0.9/ 74.8) | 177 (0.4/ 29.8) | 416 (0.8/ 70.2) |
| **Education level** |  |  | <0.001 |  |  |  |  |
| Primary education | 366 (2.8, 19.4) | 1,519 (1.5, 80.6) |  | 200 (4.1/ 54.6) | 166 (2.0/ 45.4) | 749 (1.6/ 49.3) | 770 (1.4/ 50.7) |
| Secondary/post-secondary education | 5,593 (42.6, 14.8) | 32,136 (32.1, 85.2) |  | 2,429 (49.7/ 43.4) | 3,164 (38.4/ 56.6) | 15,476 (33.5/ 48.2) | 16,660 (30.9/ 51.8) |
| Tertiary/bachelor | 4,786 (36.4, 11.7) | 36,015 (36.0, 88.3) |  | 1,443 (29.5/ 30.2) | 3,343 (40.5/ 69.8) | 14,012 (30.3/ 38.9) | 22,003 (40.8/ 61.1) |
| Master/doctoral | 2,174 (16.6, 7.1) | 28,564 (28.5, 92.9) |  | 718 (14.7/ 33.0) | 1,456 (17.7/ 67.0) | 15,040 (32.7/ 52.7) | 13,524 (25.1/ 47.3) |
| Unknown/missing/others | 216 (1.6, 10.4) | 1,853 (1.9, 89.6) |  | 99 (2.0/ 45.8) | 117 (1.4/ 54.2) | 899 (1.9/ 48.5) | 954 (1.8/ 51.5) |
| **Marital status** |  |  | <0.001 |  |  |  |  |
| Single | 4,457 (33.9, 16.5) | 22,586 (22.6, 83.5) |  | 1,348 (27.6/ 30.3) | 3,109 (37.7/ 69.7) | 10,330 (22.4/ 45.7) | 12,256 (22.7/ 54.3) |
| Married/ Civil partnership | 6,752 (51.4, 9.6) | 63,801 (63.7, 90.4) |  | 2,814 (57.6/ 41.7) | 3,938 (47.7/ 58.3) | 30,691 (66.5/ 48.1) | 33,110 (61.4/ 51.9) |
| Divorced/ Separated | 1,490 (11.4, 13.4) | 9,657 (9.6, 86.6) |  | 551 (11.3/ 37.0) | 939 (11.4/ 63.0) | 3,693 (8.0/ 38.2) | 5,964 (11.1/ 61.8) |
| Widowed | 161 (1.2, 7.3) | 2,046 (2.1, 92.7) |  | 43 (0.9/ 26.7) | 118 (1.4/ 73.3) | 476 (1.0/ 23.3) | 1,570 (2.9/ 76.7) |
| Missing | 275 (2.1, 12.1) | 1,997 (2.0, 87.9) |  | 133 (2.6/ 48.4) | 142 (1.8/ 51.6) | 986 (2.1/ 49.4) | 1,011 (1.9/ 50.6) |
| **Origin** |  |  | <0.001 |  |  |  |  |
| French by birth | 12,438 (94.7, 11.8) | 93,038 (93.0, 88.2) |  | 4,587 (93.8/ 36.9) | 7,851 (95.2/ 63.1) | 42,904 (92.9/ 46.1) | 50,134 (93.0/ 53.9) |
| France by nationality | 296 (2.2, 7.9) | 3,440 (3.4, 92.1) |  | 132 (2.7/ 44.6) | 164 (2.0/ 55.4) | 1,563 (3.4/ 45.4) | 1,877 (3.5/ 54.6) |
| Foreigner | 231 (1.8, 9.4) | 2,225 (2.2, 90.6) |  | 93 (1.9/ 40.3) | 138 (1.7/ 59.7) | 10,66 (2.3/ 47.9) | 1,159 (2.1/ 52.1) |
| Missing | 170 (1.3, 10.9) | 1,384 (1.4, 89.1) |  | 77 (1.6/ 45.3) | 93 (1.1/ 54.7) | 643 (1.4/ 46.5) | 741 (1.4/ 53.5) |
| **Household monthly average net income** |  |  | <0.001 |  |  |  |  |
| Less than 1500 € | 1,603 (12.2, 19.8) | 6,476 (6.5, 80.2) |  | 471 (9.6/ 29.4) | 1,132 (13.7/ 70.6) | 2,593 (5.6/ 40.0) | 3,883 (7.2/ 60.0) |
| 1500 € to less than 2100 € | 1,629 (12.4, 15.4) | 8,948 (8.9, 84.6) |  | 569 (11.6/ 34.9) | 1,060 (12.9/ 65.1) | 3,506 (7.6/ 39.2) | 5,442 (10.1/ 60.8) |
| 2100 € to less than 2800 € | 2,119 (16.1, 13.5) | 13,529 (13.5, 86.5) |  | 769 (15.7/ 36.3) | 1,350 (16.4/ 63.7) | 5,787 (12.5/ 42.8) | 7,742 (14.4/ 57.2) |
| 2800€ to 4200€ | 4,402 (33.5, 12.5) | 30,758 (30.7, 87.5) |  | 1,745 (35.7/ 39.6) | 2,657 (32.2/ 60.4) | 14,388 (31.2/ 46.8) | 16,370 (30.4/ 53.2) |
| More than 4200€ | 2,451 (18.7, 6.8) | 33,856 (33.8, 93.2) |  | 1,040 (21.3/ 42.4) | 1,411 (17.1/ 57.6) | 17,406 (37.7/ 51.4) | 16,450 (30.5/ 48.6) |
| Unknown | 931 (7.1, 12.5) | 6,520 (6.6, 87.5) |  | 295 (6.1/ 31.7) | 636 (7.7/ 68.3) | 2,496 (5.4/ 38.3) | 4,024 (7.5/ 61.7) |
| **Occupational level** |  |  | <0.001 |  |  |  |  |
| Labour, semi-skilled worker | 421 (3.2, 22.0) | 1,494 (1.5, 78.0) |  | 286 (5.8/ 67.9) | 135 (1.6/ 32.1) | 1,087 (2.4/ 72.8) | 407 (0.8/ 27.2) |
| Skilled worker, highly skilled worker, shop technician | 1,137 (8.7, 22.6) | 3,896 (3.9, 77.4) |  | 804 (16.4/ 70.7) | 333 (4.0/ 29.3) | 3,087 (6.7/ 79.2) | 809 (1.5/ 20.8) |
| Supervisor | 824 (6.3, 16.9) | 4,061 (4.1, 83.1) |  | 439 (9.0/ 53.3) | 385 (4.7/ 46.7) | 2,301 (5.0/ 56.7) | 1,760 (3.3/ 43.3) |
| Chief executive officer, deputy CEO | 169 (1.3, 7.8) | 2,001 (2.0, 92.2) |  | 93 (1.9/ 55.0) | 76 (0.9/ 45.0) | 1,415 (3.1/ 70.7) | 586 (1.1/ 29.3) |
| Technician, draughtsman, sales representative | 600 (4.6, 13.6) | 3,826 (3.8, 86.4) |  | 330 (6.7/ 55.0) | 270 (3.3/ 45.0) | 2,522 (5.5/ 65.9) | 1,304 (2.4/ 34.1) |
| Primary school teacher, social worker | 1,227 (9.3, 14.3) | 7,367 (7.4, 85.7) |  | 203 (4.2/ 16.5) | 1,024 (12.4/ 83.5) | 1,535 (3.3/ 20.8) | 5,832 (10.8/ 79.1) |
| Engineer, executive | 1,729 (13.2, 8.2) | 19,345 (19.3, 91.8) |  | 878 (18.0/ 50.8) | 851 (10.3/ 49.2) | 11,894 (25.8/ 61.5) | 7,451 (13.8/ 38.5) |
| Teacher, public service category personnel | 967 (7.4, 7.6) | 11,706 (11.7, 92.4) |  | 265 (5.4/ 27.4) | 702 (8.5/ 72.6) | 4,350 (9.4/ 37.1) | 7,380 (13.7/ 62.9) |
| Office or commercial employee | 2,878 (21.8, 21.3) | 10,532 (10.6, 78.7) |  | 461 (9.4/ 16.0) | 2,417 (29.4/ 84.0) | 2,179 (4.7/ 20.5) | 8,461 (15.6/ 79.5) |
| Others, missing | 3,183 (24.1, 8.2) | 35,119 (35.7, 91.8) |  | 1,130 (23.2/ 35.5) | 2,053 (24.9/ 64.5) | 15,806 (34.1/ 44.2) | 19,921 (37.0/ 55.8) |
| **Physical activity intensity (from 0 to 6)** |  |  |  |  |  |  |  |
| **Mean (SD)** | 3.4 (1.5) | 3.6 (1.5) |  | 3.5 (1.5) | 3.37 (1.4) | 3.57 (1.5) | 3.6 (1.5) |

a. *P*-value compared tattooed/ non-tattooed.

### Medical data

As part of Constances, medical data is available through the System National de Donnees de Sante (SNDS), covering all medical acts received and (partially) reimbursed by the French National Health Insurance. These data covering visits to private practices, hospitalisations, care received for chronic diseases, and drug prescriptions are complemented by self-reported medical questionnaires. At baseline, current physical and psychological well-being and lifetime medical history were assessed. Annual follow-ups capture additional medical information such as physical and psychological well-being, as well as occurrence of specific diagnoses including incident cancer diagnoses during the 12 months prior. Furthermore, individual case verification via recontact of incidence cancer cases can provide more detailed information on cancer diagnosis.

## Results

Out of 112,222 Constances study participants who answered the follow-up questionnaire in 2020, 13,135 (11.7%) reported having at least one tattoo (supplementary figure 1). Full exposure data assessed via the EpiTAT exposure questionnaire in 2023 was collected from 7,928 tattooed participants (response rate of 60.4%). The remaining 5,207 (39.6%) constitute the group of tattooed non-responders in the present analysis. We excluded 37 (0.3%) surveys which were returned without answers.

Table 1 shows various demographic and lifestyle characteristics of individuals categorised by tattoo status and sex. Overall, the study population consists of more women than men, and even more so among those with tattoos (women: n=8,246 (62.8%); men: n=4,889 (37.2%)). While in the total cohort the older age groups predominate, tattoo prevalence decreases with increasing age leading to an inverse age/tattoo distribution within the cohort.

Table 1 shows strong sociodemographic and lifestyle differences in between the tattooed and noin-tattooed cohort population. Among both sexes, tattooed individuals had a higher prevalence of substance use compared to non-tattooed individuals. Current smoking status, alcohol abuse and dependence, ever use of e-cigarettes and ever use of cannabis were all higher in tattooed compared to non-tattooed individuals. Patterns were similar when stratified by sex. Tattooed compared to non-tattooed individuals also had slightly higher percentages of underweight and obese groups, had a lower income, were lower educated, and less often in a romantic relationship.

### Tattoo exposure characteristics

To assess tattooing-incurred cancer risk, profound knowledge on the exposure is key: tattoo pigment toxicity varies by tattoo colour, and due to different forms and shapes of tattoos, the tattoo “filling” needs to be considered when estimating the total tattooed body surface. Furthermore, the time since tattooing defines the assumed lag-time to cancer formation and the tattoo circumstances give information about potential infections related to tattooing that potentially mediate the tattoo-cancer relationship. Table 2 provides a comprehensive overview of tattoo characteristics for individuals with known exposure details. The median tattoo size of tattooed participants was one hand surface (interquartile range (IQR)= 0.5; 3.0)) and translated to a metric median of 217.6 cm^2^ (94.7; 536.8). Tattoo size was highest in younger age groups and in men (Figure 1). By adjusting for tattoo filling, the median tattoo surface area among overall tattooed participants decreased to nearly half its original size.

**Table 2:**
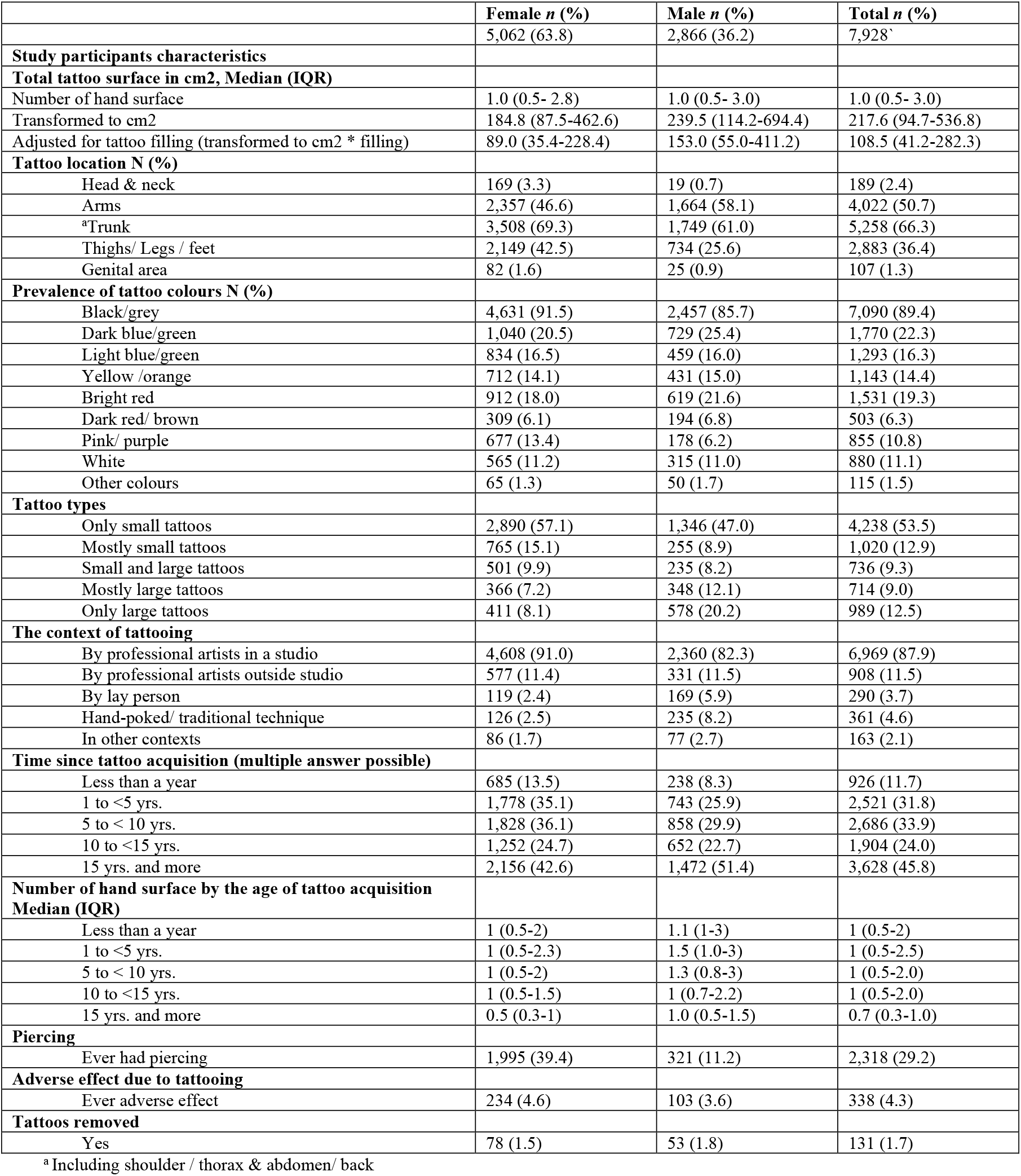
Tattoo characteristics amongst tattooed individuals with full exposure information.

|  | Female n (%) | Male n (%) | Total n (%) |
| --- | --- | --- | --- |
|  | 5,062 (63.8) | 2,866 (36.2) | 7,928<sup>a</sup> |
| **Study participants characteristics** |  |  |  |
| **Total tattoo surface in cm2, Median (IQR)** |  |  |  |
| Number of hand surface | 1.0 (0.5- 2.8) | 1.0 (0.5- 3.0) | 1.0 (0.5- 3.0) |
| Transformed to cm2 | 184.8 (87.5-462.6) | 239.5 (114.2-694.4) | 217.6 (94.7-536.8) |
| Adjusted for tattoo filling (transformed to cm2 * filling) | 89.0 (35.4-228.4) | 153.0 (55.0-411.2) | 108.5 (41.2-282.3) |
| **Tattoo location N (%)** |  |  |  |
| Head & neck | 169 (3.3) | 19 (0.7) | 189 (2.4) |
| Arms | 2,357 (46.6) | 1,664 (58.1) | 4,022 (50.7) |
| <sup>a</sup>Trunk | 3,508 (69.3) | 1,749 (61.0) | 5,258 (66.3) |
| Thighs/ Legs / feet | 2,149 (42.5) | 734 (25.6) | 2,883 (36.4) |
| Genital area | 82 (1.6) | 25 (0.9) | 107 (1.3) |
| **Prevalence of tattoo colours N (%)** |  |  |  |
| Black/grey | 4,631 (91.5) | 2,457 (85.7) | 7,090 (89.4) |
| Dark blue/green | 1,040 (20.5) | 729 (25.4) | 1,770 (22.3) |
| Light blue/green | 834 (16.5) | 459 (16.0) | 1,293 (16.3) |
| Yellow /orange | 712 (14.1) | 431 (15.0) | 1,143 (14.4) |
| Bright red | 912 (18.0) | 619 (21.6) | 1,531 (19.3) |
| Dark red/ brown | 309 (6.1) | 194 (6.8) | 503 (6.3) |
| Pink/ purple | 677 (13.4) | 178 (6.2) | 855 (10.8) |
| White | 565 (11.2) | 315 (11.0) | 880 (11.1) |
| Other colours | 65 (1.3) | 50 (1.7) | 115 (1.5) |
| **Tattoo types** |  |  |  |
| Only small tattoos | 2,890 (57.1) | 1,346 (47.0) | 4,238 (53.5) |
| Mostly small tattoos | 765 (15.1) | 255 (8.9) | 1,020 (12.9) |
| Small and large tattoos | 501 (9.9) | 235 (8.2) | 736 (9.3) |
| Mostly large tattoos | 366 (7.2) | 348 (12.1) | 714 (9.0) |
| Only large tattoos | 411 (8.1) | 578 (20.2) | 989 (12.5) |
| **The context of tattooing** |  |  |  |
| By professional artists in a studio | 4,608 (91.0) | 2,360 (82.3) | 6,969 (87.9) |
| By professional artists outside studio | 577 (11.4) | 331 (11.5) | 908 (11.5) |
| By lay person | 119 (2.4) | 169 (5.9) | 290 (3.7) |
| Hand-poked/ traditional technique | 126 (2.5) | 235 (8.2) | 361 (4.6) |
| In other contexts | 86 (1.7) | 77 (2.7) | 163 (2.1) |
| **Time since tattoo acquisition (multiple answer possible)** |  |  |  |
| Less than a year | 685 (13.5) | 238 (8.3) | 926 (11.7) |
| 1 to <5 yrs. | 1,778 (35.1) | 743 (25.9) | 2,521 (31.8) |
| 5 to < 10 yrs. | 1,828 (36.1) | 858 (29.9) | 2,686 (33.9) |
| 10 to <15 yrs. | 1,252 (24.7) | 652 (22.7) | 1,904 (24.0) |
| 15 yrs. and more | 2,156 (42.6) | 1,472 (51.4) | 3,628 (45.8) |
| **Number of hand surface by the age of tattoo acquisition Median (IQR)** |  |  |  |
| Less than a year | 1 (0.5-2) | 1.1 (1-3) | 1 (0.5-2) |
| 1 to <5 yrs. | 1 (0.5-2.3) | 1.5 (1.0-3) | 1 (0.5-2.5) |
| 5 to < 10 yrs. | 1 (0.5-2) | 1.3 (0.8-3) | 1 (0.5-2.0) |
| 10 to <15 yrs. | 1 (0.5-1.5) | 1 (0.7-2.2) | 1 (0.5-2.0) |
| 15 yrs. and more | 0.5 (0.3-1) | 1.0 (0.5-1.5) | 0.7 (0.3-1.0) |
| **Piercing** |  |  |  |
| Ever had piercing | 1,995 (39.4) | 321 (11.2) | 2,318 (29.2) |
| **Adverse effect due to tattooing** |  |  |  |
| Ever adverse effect | 234 (4.6) | 103 (3.6) | 338 (4.3) |
| **Tattoos removed** |  |  |  |
| Yes | 78 (1.5) | 53 (1.8) | 131 (1.7) |

<sup>a</sup> Including shoulder / thorax & abdomen/ back

**Figure 1:**
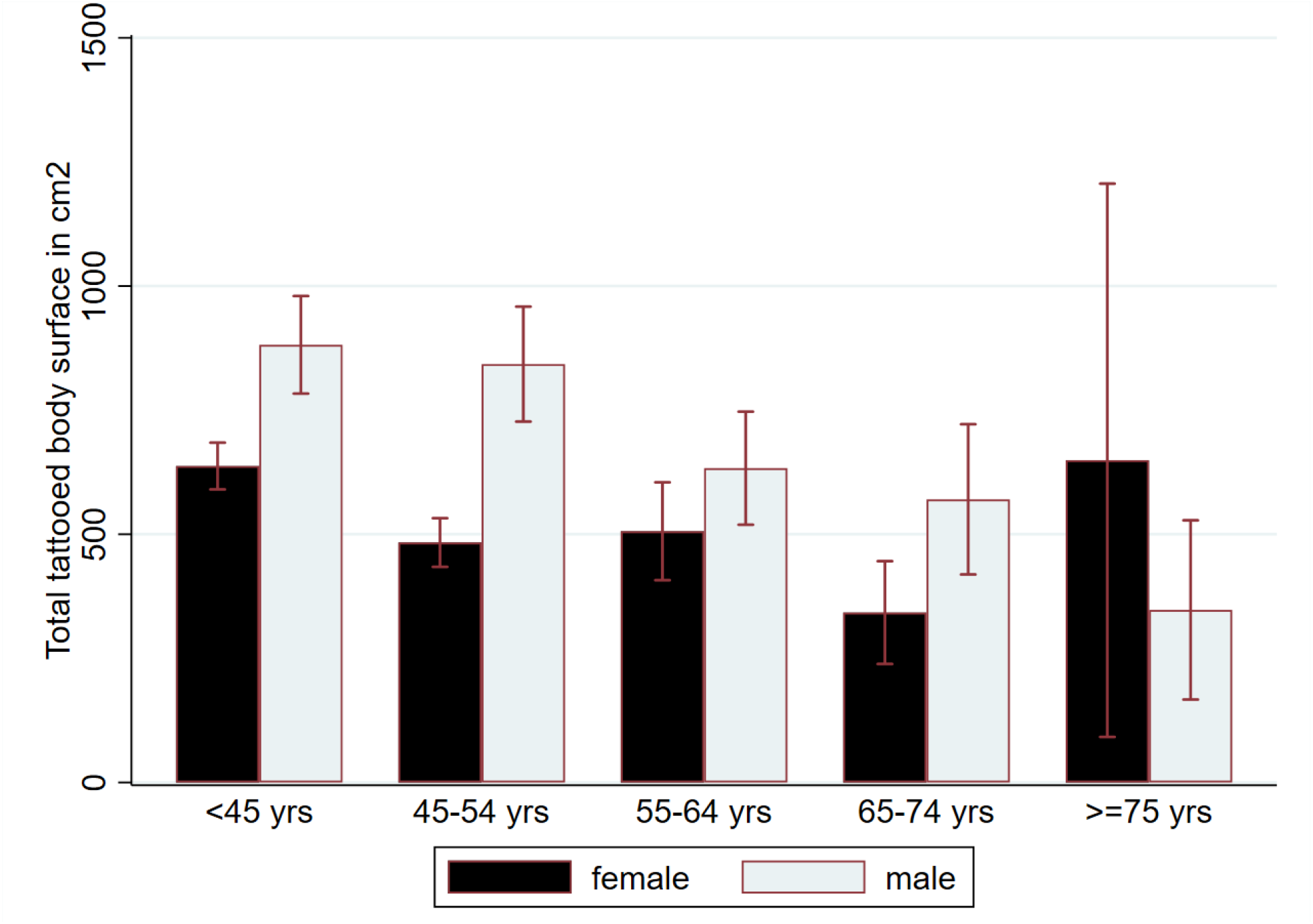
The mean total body surface tattooed by biological sex over age categories in the tattooed subsample with full exposure information.

The location of tattoos varied by sex. The trunk (including shoulders, back and abdomen) was the most common tattooed area, with approximately 70% of females and 60% of males having tattoos on this body part. This was followed by the arms, which were tattooed more often by males than females, while women had more tattoos on the legs and feet than males.

By far, the most common tattoo colour was black/grey. Other common colours reported by 15-20% of tattooed participants were dark blue/green, bright red and light blue/green and yellow/orange. Other colours such as pink/violet, white and dark red/brown were less than 10% in total (Figure 2).

**Figure 2:**
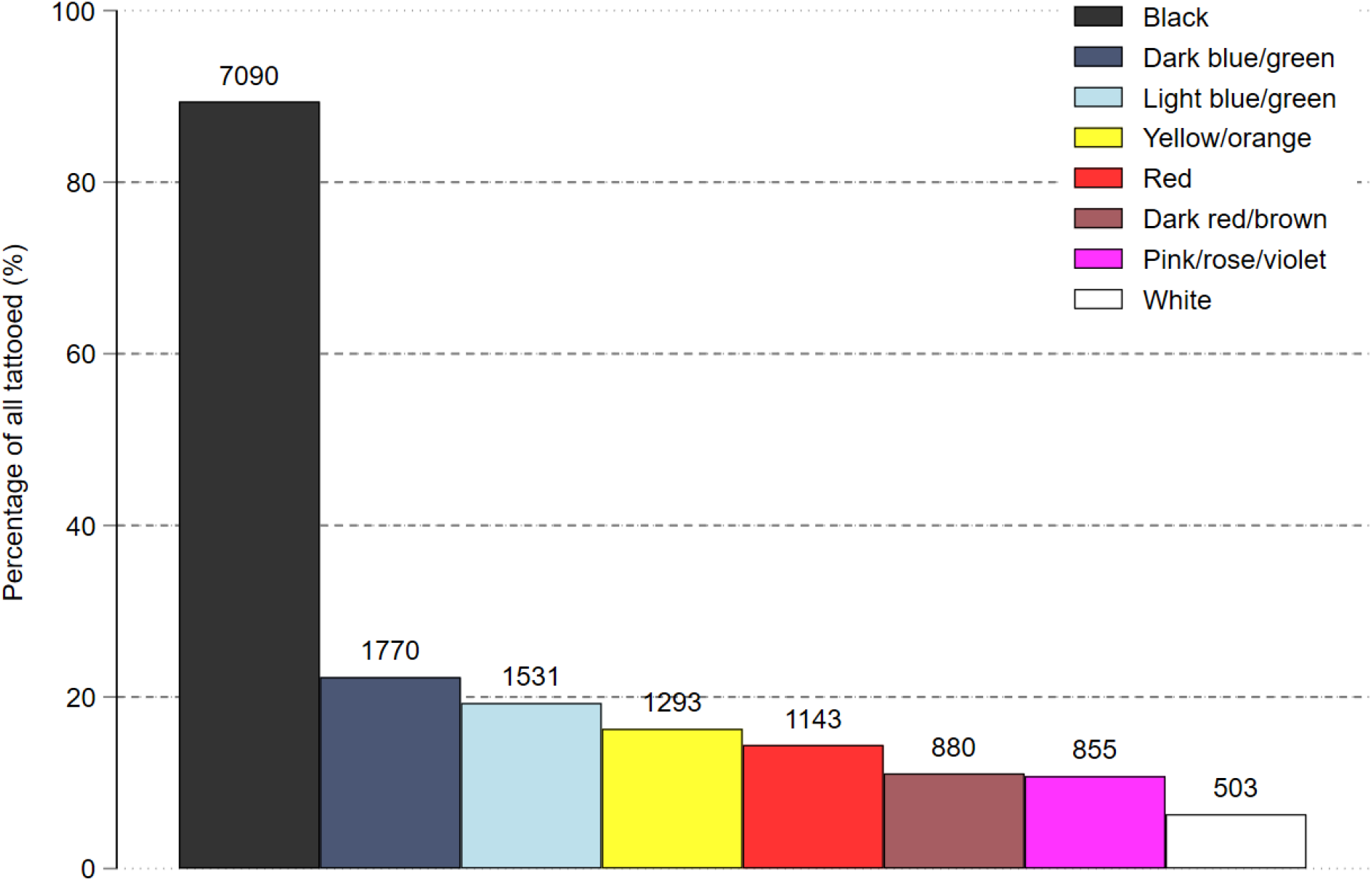
Frequency and percentage of tattoo colours in the tattooed subsample with full exposure information.

In term of tattoo sizes, about half of participants had only small tattoos, whilst one fifth had mostly or only large. Females were more likely to prefer small and men more likely to prefer large tattoos.

Almost 9 in 10 tattooed individuals received at least one tattoo in professional studios, while more than one in 10 also were tattooed outside of studios by professionals. Together with tattoos acquired from laypersons, and through traditional or hand-poked techniques, about one fifth of tattooed individuals had at least one tattoo from outside a studio (data not shown).

Almost half of participants had tattoos for more than 15 years with a higher proportion of males than females. Tattoos acquired within 5-10 years were slightly more common among females than males. More than one out of 10 tattooed individuals reported a new tattoo during the last year. About one third of participants had piercings.

Adverse effects from tattooing occurred less than 5%, and were slightly more often reported in females than males. Furthermore, when it came to protecting their tattoos from the sun, 36.2% sometimes protected their tattoos. Females were more likely to always protect their tattoos from the sun than males.

Less than 2% underwent tattoos removal, thereby laser removal was most common, followed by dermabrasion. The size of the removal was usually less than the area of a hand and more than 10% reported adverse effects of removal.

Further results on the prevalence of tattoo acquisition outside of France by tattooed participants showed, while Europe (neighbouring countries including Spain, United Kingdom, Belgium, Portugal, and Germany) and Asia (Thailand), and North America, and Canada are the most common regions outside of France for tattoo acquisition among participants, fewer participants obtained tattoos from South America and Africa (data not shown).

### Comparison with non-responders

As seen in Table 3, the sociodemographic profile of tattooed participants who responded differed slightly from participants who did not respond to the EpiTAT questionnaire. For example, compared to non-respondents, the respondents more often had a university degree, were married or in a civil partnership, and had higher incomes.

**Table 3:** Participant characteristic of tattooed individuals who either responded or did not respond to the EpiTAT questionnaire.

|  | **Tattooed/**<br>**Responded** | **Tattooed/**<br>**No respond** | **P-value\*** |
| --- | --- | --- | --- |
| N (%) | 7,928 (7.0) | 5,207 (4.6) |  |
| **Sex** |  |  | 0.002 |
| Male | 2,866 (36.2) | 2,023 (38.9) |  |
| Female | 5,062 (63.8) | 3,184 (61.1) |  |
| **Age** |  |  | 0.039 |
| <45 yrs. | 3,231 (40.7) | 2,203 (42.3) |  |
| 45-54 yrs. | 2,368 (29.9) | 1,439 (27.6) |  |
| 55-64 yrs. | 1,514 (19.1) | 1,017 (19.5) |  |
| 65-74 yrs. | 707 (8.9) | 491 (9.5) |  |
| >=75 yrs. | 108 (1.4) | 57 (1.1) |  |
| **Tobacco smoking** |  |  | <0.001 |
| Never | 2,848 (35.9) | 1,577 (30.3) |  |
| Former | 2,914 (36.7) | 1,803 (34.6) |  |
| Current | 1,886 (23.8) | 1,573 (30.2) |  |
| Missing | 280 (3.6) | 254 (4.9) |  |
| **Alcohol consumption** |  |  | <0.001 |
| No drinker | 143 (1.8) | 124 (2.4) |  |
| Current drinker (abuse) | 1,588 (20.0) | 1,059 (20.3) |  |
| Current drinker (dependent) | 437 (5.5) | 369 (7.1) |  |
| Current drinker (not dependent) | 5,265 (66.5) | 3,264 (62.7) |  |
| Unknown | 495 (6.2) | 391 (7.5) |  |
| **E-cigarette** |  |  | <0.001 |
| Never | 4,908 (61.9) | 3,130 (60.1) |  |
| Ever | 1,049 (13.2) | 910 (17.5) |  |
| Missing | 1,971 (24.9) | 1,167 (22.4) |  |
| **Cannabis consumption** |  |  | <0.001 |
| Never | 3,339 (42.1) | 2,056 (39.5) |  |
| Ever | 4,364 (55.1) | 2,914 (56.0) |  |
| Missing | 225 (2.8) | 237 (4.5) |  |
| **Body Mass Index kg/M<sup>2</sup>** |  |  | 0.032 |
| <18.5 (underweight) | 253 (3.2) | 170 (3.3) |  |
| 18.5-24.9 (healthy range) | 4,556 (57.5) | 2,848 (54.7) |  |
| 25-29.9 (overweight) | 2,196 (27.6) | 1,508 (29.0) |  |
| 30-39.9 (obesity) | 861 (10.9) | 640 (12.2) |  |
| >40 (severe obesity) | 62 (0.8) | 41 (0.8) |  |
| **Education level** |  |  | <0.001 |
| Early childhood/primary education | 178 (2.2) | 188 (3.6) |  |
| Secondary/post-secondary education | 3,121 (39.4) | 2,472 (47.5) |  |
| Tertiary/bachelor | 3,076 (38.8) | 1,710 (32.8) |  |
| Master/doctoral | 1,456 (18.4) | 718 (13.8) |  |
| Unknown/missing/others | 97 (1.2) | 119 (2.3) |  |
| **Marital status** |  |  | 0.002 |
| Single | 2,621 (33.1) | 1,836 (35.2) |  |
| Married/ Civil partnership | 4,200 (53.0) | 2,552 (49.0) |  |
| Divorced/ Separated | 896 (11.3) | 594 (11.4) |  |
| Widow | 90 (1.1) | 71 (1.4) |  |
| Missing | 121 (1.5) | 154 (3.0) |  |
| **Origin** |  |  | <0.001 |
| French by birth | 7,578 (95.6) | 4,860 (93.3) |  |
| France by nationality | 153 (1.9) | 143 (2.7) |  |
| Foreigner | 113 (1.4) | 118 (2.3) |  |
| Missing | 84 (1.1) | 86 (1.7) |  |
| **Household average net income** |  |  | <0.001 |
| less than 1500 € | 850 (10.7) | 753 (14.5) |  |
| 1500 € to less than 2100 € | 950 (12.0) | 679 (13.0) |  |
| 2100 € to less than 2800 € | 1,274 (16.1) | 845 (16.2) |  |
| 2800€ to 4200€ | 2,740 (34.5) | 1,662 (31.9) |  |
| More than 4200€ | 1,614 (20.4) | 837 (16.1) |  |
| Unknown | 500 (6.3) | 431 (8.3) |  |
| **Occupational level** |  |  | <0.001 |
| Labour, semi-skilled worker | 215 (2.7) | 206 (4.0) |  |
| Skilled worker, highly skilled worker, shop technician | 629(7.9) | 508 (9.8) |  |
| Supervisor | 497 (6.3) | 327 (6.3) |  |
| Chief executive officer, deputy ceo | 91 (1.1) | 78 (1.5) |  |
| Technician, draughtsman, sales representative | 397 (5.0) | 203 (3.9) |  |
| Primary school teacher, social worker | 801 (10.1) | 426 (8.2) |  |
| Engineer, executive | 1,138 (14.4) | 591 (11.4) |  |
| Teacher, public service category personnel | 664 (8.4) | 303 (5.8) |  |
| office or commercial employee | 1,706 (21.5) | 1,172 (22.5) |  |
| Others, missing | 1,790 (22.6) | 1,393 (26.8) |  |

\*p-value compared tattooed responded/non-responded
\*\* Unknown/missing refers to individuals who did not respond to the question of whether they had tattoos in 2020.

## Discussion

Comprised of over 13,000 tattooed participants and 100,000 non-tattooed controls, CRABAT is, to our knowledge, the largest cohort study on tattoo-associated long-term health effects worldwide. As a main strength, CRABAT is nested in the French national Constances cohort, providing rich complementary data and allowing for long-term prospective follow-up. Its individual record linkage to the national health insurance assures objective health outcome data. In using the Constances infrastructure, CRABAT can address a multitude of research questions in selecting for each of them separately the relevant outcome and covariate data from the study variable pool. The strong sociodemographic differences of the tattooed compared to the non-tattooed study population, particularly for known cancer risk factors, underscores the importance of the available complementary data to consider in risk analyses. Moreover, the exceptional tattoo exposure data that was collected using a validated questionnaire in almost 8,000 participants, will allow for assessment of dose-response relationships and stratification on visual or contextual tattoo factors.

As a weakness of many cohort studies, CRABAT relies on self-reported data. As an example, the validation study of the EpiTAT exposure questionnaire showed that self-reported tattoo size is strongly overestimated (17). Visual mobile phone assisted tattoo surface measures might become a more accurate alternative but were not available at the time of data collection. In addition, the response rate to the EpiTAT exposure questionnaire was 60.2%, which reduced the tattooed sample with detailed exposure characteristics. While the complexity of the EpiTAT questionnaire could have discouraged some participants, the non-responders also seemed to be generally from less privileged social classes typical in many studies. To eliminate potential non-responder bias (i.e. if they systematically differ from responders on key characteristics of exposure such as the size of tattoos) and to catch newly tattooed participants, we plan a second wave of basic exposure assessment in 2030. Finally, owing to its design and presenting an asset and limitation at once, the Constances cohort population is a particularly healthy population as it consists of volunteers that agreed to participate in a life-long cohort. We can assume that the tattoo prevalence in the general population is higher than in Constances, and also that known cancer risk factors (e.g. smoking, alcohol abuse) could be more common in the general population than in Constances. While this may reduce the generalizability of the (tattooed) Constances population for the (tattooed) population as a whole, estimation of cancer risks might be less confounded by confounding factors that predominantly affect tattooed individuals.

## Data access

Due to European data protection regulations, the CRABAT data cannot be made available to the public. However, access to the data may be granted via the standard project application procedure to the Constances infrastructure and subsequent approval by the Institutional Steering Committee. More information on the application procedure and the terms of use can be found on the Consatnces website: https://www.constances.fr/index_EN.

## Ethics approval

The CRABAT study received additional approval by the IARC Ethics Committee (IEC 22-02), and was authorized by the CNIL (#22015584).The Constances study was approved by the Institutional review board (IRB) of the French Institute of Health (Inserm) (Opinion n°01-011, then n°21-842), and authorized by the by the French Data Protection Authority (“Commission Nationale de l’Informatique et des Libertés”, CNIL) (Authorization #910486).

## Data availability

Data are not publicly available and can be obtained from third parties. Access to the data is subject to the submission of a research project proposal, which will be reviewed by the CONSTANCES International Scientific Committee and must be approved by the Institutional Steering Committee.

## Supplementary data

Supplementary data are available at *IJE* online.

## Author contributions

M.F., J.S., I.S. had substantial contributions to the design and conduct of the work. B.H., R.D.M., M.F., drafted and prepared the manuscript. B.H., and M.F., conducted the statistical analyses. The manuscript was reviewed and commented on by all authors. The final version of the manuscript has been approved by all authors. The remaining authors (M.Z., M.G., S.K., C.R., K.E.) contributed to the main cohort study and ensured the quality of the original data and ensuring that questions related to the accuracy of any part of the work are appropriately investigated. All authors contributed to the interpretation of the results, revising it critically for important intellectual content. M.F., contributors as being responsible for the overall content as guarantors. The work reported in the paper has been performed by the authors, unless clearly specified in the text.

## Disclaimer

Where authors are identified as personnel of the International Agency for Research on Cancer/World Health Organization, the authors alone are responsible for the views expressed in this article and they do not necessarily represent the decisions, policy, or views of International Agency for Research on Cancer /World Health Organization

## References

1. Kluger N. Epidemiology of tattoos in industrialized countries. Tattooed skin and health: Karger Publishers; 2015. p. 6–20.

2. Giulbudagian M, Schreiver I, Singh AV, Laux P, Luch A. Safety of tattoos and permanent make-up: a regulatory view. Archives of Toxicology. 2020;94:357–69.

3. Bäumler W. Chemical hazard of tattoo colorants. La Presse Médicale. 2020;49(4):104046.

4. Kürle S, Schulte KW, Homey B. [Accumulation of tattoo pigment in sentinel lymph nodes]. Hautarzt. 2009;60(10):781–3. Epub 2009/09/17. doi: 10.1007/s00105-009-1843-9. PubMed PMID: 19756437.

5. Loomis D, Guha N, Hall AL, Straif K. Identifying occupational carcinogens: an update from the IARC Monographs. Occupational and environmental medicine. 2018;75(8):593–603.

6. DeMarini DM, Carreón-Valencia T, Gwinn WM, Hopf NB, Sandy MS, Bahadori T, Calaf GM, Chen G, de Conti A, Fritschi L. Carcinogenicity of some aromatic amines and related compounds. The Lancet Oncology. 2020;21(8):1017–8.

7. Baan R, Grosse Y, Straif K, Secretan B, El Ghissassi F, Bouvard V, Benbrahim-Tallaa L, Guha N, Freeman C, Galichet L. A review of human carcinogens—part F: chemical agents and related occupations. The lancet oncology. 2009;10(12):1143–4.

8. Schreiver I, Hesse B, Seim C, Castillo-Michel H, Villanova J, Laux P, Dreiack N, Penning R, Tucoulou R, Cotte M. Synchrotron-based ?-XRF mapping and μ-FTIR microscopy enable to look into the fate and effects of tattoo pigments in human skin. Scientific reports. 2017;7(1):11395.

9. Foerster M, Schreiver I, Luch A, Schüz J. Tattoo inks and cancer. Cancer Epidemiology. 2020;65.

10. Serup J, Carlsen KH, Sepehri M. Tattoo complaints and complications: diagnosis and clinical spectrum. Curr Probl Dermatol. 2015;48:48–60. Epub 2015/04/04. doi: 10.1159/000369645. PubMed PMID: 25833625.

11. Barton DT, Zens MS, Marmarelis EL, Gilbert-Diamond D, Karagas MR. Cosmetic Tattooing and Early Onset Basal Cell Carcinoma: A Population-based Case–Control Study from New Hampshire. Epidemiology. 2020;31(3):448–50.

12. Nielsen C, Jerkeman M, Jöud AS. Tattoos as a risk factor for malignant lymphoma: a population-based case–control study. EClinicalMedicine. 2024;72.

13. McCarty RD, Trabert B, Millar MM, Haaland B, Grieshober L, Barnard M, Collin L, Gilreath JA, Shami PJ, Doherty JA. Tattooing and risk of hematologic cancer: A population-based case-control study in Utah. Cancer Research. 2023;83(7_Supplement):6471-.

14. Warner FM, Darvishian M, Boyle T, Brooks-Wilson AR, Connors JM, Lai AS, L. ND, Song K, Sutherland H, Woods RR. Tattoos and Hematologic Malignancies in British Columbia, Canada. Cancer Epidemiology and Prevention Biomarkers. 2020;29(10):2093–5.

15. Goldberg M, Carton M, Descatha A, Leclerc A, Roquelaure Y, Santin G, Zins M. CONSTANCES: a general prospective population-based cohort for occupational and environmental epidemiology: cohort profile. Occupational and Environmental Medicine. 2016.

16. Siegrist J, Goldberg M, Zins M, Wahrendorf M. Social inequalities, stressful work and non-fatal cardiovascular disease: follow-up findings from the CONSTANCES Study. Occupational and Environmental Medicine. 2023.

17. Foerster M, Dufour L, Bäumler W, Schreiver I, Goldberg M, Zins M, Ezzedine K, Schüz J. Development and Validation of the Epidemiological Tattoo Assessment Tool to Assess Ink Exposure and Related Factors in Tattooed Populations for Medical Research: Cross-sectional Validation Study. JMIR Formative Research. 2023;7:e42158.

18. Sacco JJ, Botten J, Macbeth F, Bagust A, Clark P. The average body surface area of adult cancer patients in the UK: a multicentre retrospective study. PloS one. 2010;5(1):e8933.

