## Supplemental material for "Cohort profile: The Cancer Risk Associated with the Body Art of Tattooing (CRABAT) study"

### Supplementary material

*TableS1: Additional data on lifestyle factors, environmental risk factors and medical conditions available from the Constances variable pool and available in CRABAT.*

| **Variables category** | **Variables name** | **Baseline** | **Follow up** |
|---|---|---|---|
| **Lifestyle and health** | | | |
| General heath | | | 2022 |
| | Physical health | | 2014 |
| | Mental health | | 2014 |
| Physical activities | | | 2022 |
| | at work | | |
| | off work | | |
| Home and quality of life | | | |
| | Nationality | | |
| | Education level | | |
| | Marital status | | |
| | Income | | |
| Sex life | | Baseline | |
| | Number of partners | | |
| | Sextual orientation | | |
| | Use protection | | |
| Smoking | | | 2022 |
| | Lifetime tobacco smoking status | | |
| | Pack-year | | |
| Electronic cigarette | | | |
| | Lifetime smoking status | | 2022 |
| | Current smoker | | |
| | Nicotine consumption level | | |
| Cannabis consumption | | | 2022 |
| | Lifetime consumption status | | |
| | Current consumption | | |
| Alcohol consumption | | | |
| | Lifetime consumption status | | 2022 |
| | Level of consumption | | |
| **Females health** | | Baseline | |
| | Pregnancies (N) | | |
| | Date of last pregnancy | | |
| | ectopic pregnancy (N) | | |
| | Miscarriages (N) | | |
| | Therapeutic abortions (N) | | |
| | Childs (N) | | |
| | Contraception type | | 2016 |
| **Occupational history** | | | |
| | Status of employment | | 2019 |
| | Current socio-professional status | | 2019 |
| Occupational exposures | | | |
| | Night shift | | 2019 |
| Exposure to solvents/ diluent/ detergents | | Baseline | |
| | Diesel engine exhaust | | |
| | Petrol | | |
| | Trichloroethylene | | |
| | White spirit | | |
| | Cellulose diluent | | |
| | Formaldehyde | | |
| Exposure to fumes | | Baseline | |
| | fumes | | |
| | Plastic/ rubber fumes | | |
| | Biomass fumes Welding | | |
| Exposure to dusts | | Baseline | |
| *Construction dusts* | | | |
| | Asbestos | | |
| | Cement | | |
| | No-slump concrete dust | | |

| | | | |
|---|---|---|---|
| Metal dust | | Baseline | |
| | Iron | | |
| | Stainless steel | | |
| | Copper | | |
| Other dusts | | Baseline | |
| | Plastic/ rubber | | |
| | Coal | | |
| | Wood | | |
| | Textile | | |
| Fuel | | Baseline | |
| | Disel oil | | |
| | Petrol | | |
| Other chemical exposure | | | |
| | Pesticide/ insecticide/ fungicide | | |
| | Fertilizers | | |
| | Glass wood | | |
| | Insulation material | | |
| | Paint/ varnish | | |
| | Ink/ stains | | |
| | Radiation exposure | | |
| | Artificial UV radiation | | |
| | Outdoor work (hr/day) | | |
| *Biological exposure* | | Baseline | |
| | Micro-organism/ parasites/ viruses | | |
| **Medical background** | | | 2022 |
| Cardiovascular disease | | | |
| Digestive disease | | | |
| | Hepatitis B | | |
| | Hepatitis C | | |
| | Other type of hepatitis | | |
| Neurological and mental health | | | 2022 |
| | Depression | | |
| | Suicide attempts | | |
| Osteoarticular disorders | | | |
| | Inflammation arthritis | | |
| Cancer history | | | |
| Biometry | | | |
| | Body Mass Index | | 2022 |
| | Hip measure | | |
| Prothesis (N) | | | 2022 |

*Supplementary figure 1: study flowchart*

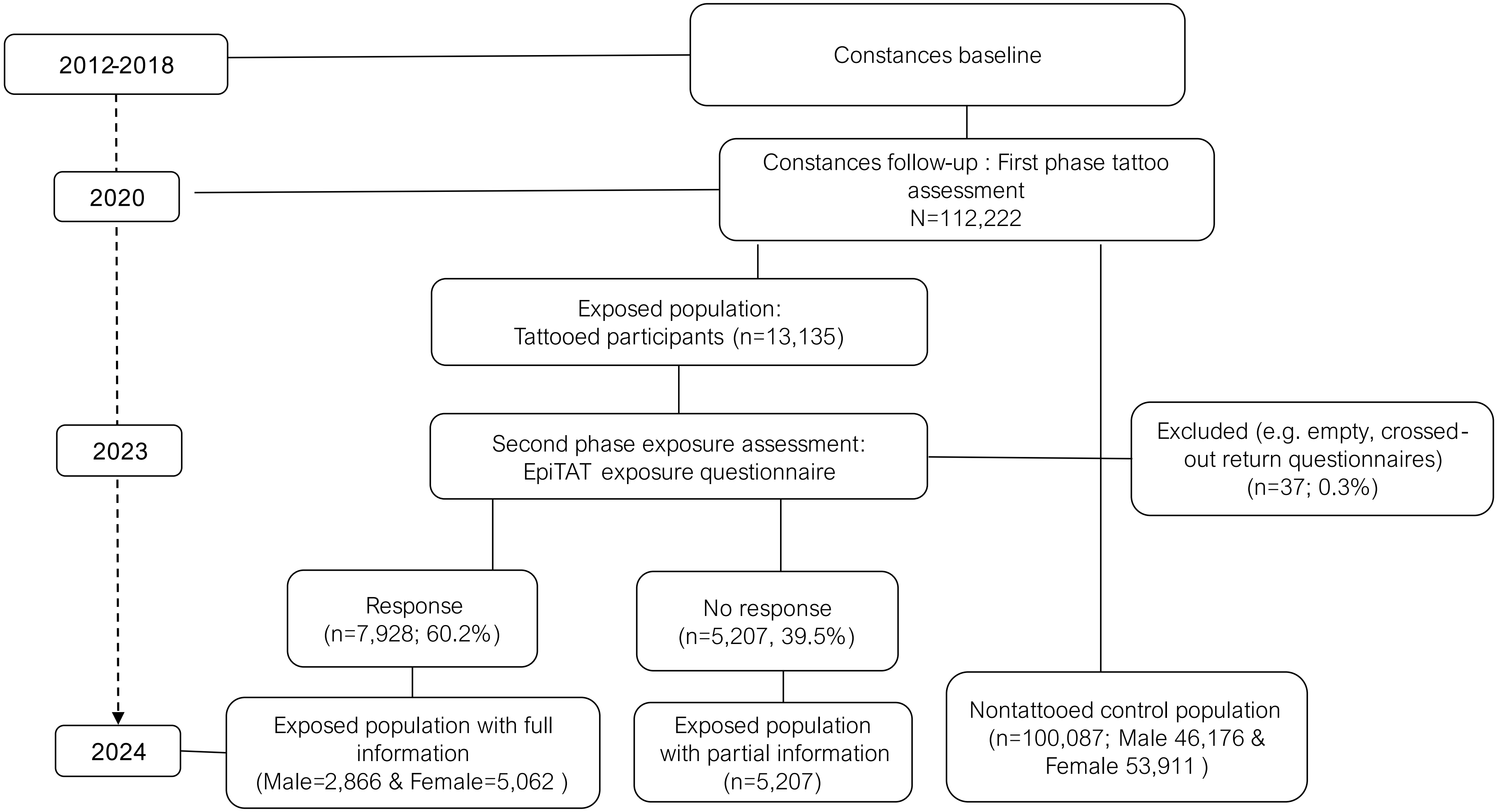
